# Cardiac Implantable Electronic Devices in Patients Referred for Transcatheter Tricuspid Edge-to-Edge Repair: Clinical Characteristics and Impact on Procedure Eligibility

**DOI:** 10.1101/2025.11.03.25339431

**Authors:** Adam Rdzanek, Adam Piasecki, Jacopo Lin, Mariusz Tomaniak, Maria Królikowska, Pavlo Matsko, Agata Markiewicz, Michał Janiel, Ewa Pędzich, Agnieszka Kapłon Cieślicka, Ewa Ostrowska, Paweł Pawłowicz, Jarosław Trębacz, Rafał Gałąska, Patrycjusz Stokłosa, Sebastian Stefaniak, Michał Chmielecki, Paolo Denti, Andrzej Gackowski, Marek Grygier, Jerzy Pręgowski, Marcin Grabowski, Piotr Scisło, Francesco Maisano

## Abstract

**Background:** Tricuspid transcatheter edge-to-edge repair (T-TEER) has become an established therapeutic option for patients with severe tricuspid regurgitation (TR). However, data regarding the qualification process for T-TEER in patients with cardiac implantable electronic devices (CIED) are limited.

**Methods:** This retrospective, multicenter study included consecutive patients with severe TR referred between January and December 2024 to six tertiary centers (five in Poland, one in Italy) for T-TEER screening. Echocardiographic assessment followed current recommendations using a five-grade TR severity scale. In CIED carriers, the relationship between the tricuspid valve and the lead was classified as CIED-related (direct interference) or CIED-associated (no causal interaction). Heart teams evaluated all cases, recording qualification status and reasons for disqualification.

**Results:** Among 271 patients (mean age 77 ± 9 years, 58.3% female), 113 (41.7%) had a CIED. Compared with non-CIED patients, CIED carriers showed higher rates of diabetes mellitus (35.4% vs. 17.1%), chronic kidney disease (70.8% vs. 54.4%), and previous CABG (13.3% vs. 4.4%), and exhibited lower LV ejection fraction (50.5% vs. 57.0%, all p < 0.05). Overall, 56.5% were qualified for T-TEER, with no significant difference between CIED and non-CIED groups (54.0% vs. 58.3%, p = 0.487). Among CIED patients, 37 (32.7%) had CIED-related TR, characterized by more severe TR (EROA 0.90 vs. 0.60 cm², p = 0.017) but similar qualification rates (59.5% vs. 52.7%, p = 0.500).

**Conclusions:** CIED carriers with severe TR represent a more complex and symptomatic population, yet the presence of a CIED does not affect eligibility for T-TEER. The high prevalence of CIED-related TR underscores the growing clinical relevance of this condition and highlights the need for prospective multicenter studies to refine patient selection and treatment strategies.

## INTRODUCTION

Transcatheter tricuspid edge-to-edge repair (T-TEER) has recently become a widely adopted treatment for patients with symptomatic tricuspid regurgitation (TR). Increasing availability of this procedure has translated into a rapid rise in the number of patients referred to tertiary centers for evaluation and consideration of transcatheter therapy.

A substantial proportion of referred patients have been previously treated with cardiac implantable electronic device (CIED) having at least one pacing or defibrillating lead crossing the tricuspid valve (TV).[1] In most cases, the presence of such a lead does not exacerbate TR severity (CIED-associated TR). However, in some patients, lead interference with the valve leaflets or subvalvular apparatus contributes significantly to the TR severity (CIED-related TR).[2] Although isolated reports suggest a potentially adverse interaction between a presence of a TV-crossing lead and T-TEER feasibility and efficacy,[3] most studies to date indicate that overall procedural outcomes of T-TEER are comparable between patients with and without CIEDs.[4,5] Nevertheless, the presence of a lead may represent a potential challenge during the intervention, hampering adequate leaflet grasping or impairing echocardiographic visualization.[6] Consequently, the presence of a CIED may influence the qualification process for T-TEER.

Although procedural outcomes of tricuspid T-TEER in patients with CIEDs have been reported, no study to date has specifically focused on the qualification process for this intervention in this challenging subset. We therefore aimed to characterize patients with and without CIEDs and to evaluate the impact of both CIED presence and the type of lead– tricuspid valve interaction (CIED-related vs. CIED-associated) on the qualification for T-TEER in a real-world, unselected cohort of patients with severe TR referred to tertiary centers.

## METHODS

### Study population

This retrospective, multicenter analysis included consecutive patients with severe TR referred between January 2024 and December 2024 to five tertiary care centers in Poland and one in Italy for screening and consideration for T-TEER.

Transthoracic and transesophageal echocardiographic examinations were performed by dedicated cardiologists with extensive experience in valvular heart disease imaging. The TV was assessed according to current guidelines using a five-grade TR severity scale based on quantitative characterization of TR.[7] In patients with CIEDs, the relationship between a TV-crossing lead and TR was determined by echocardiography. CIED-associated TR was diagnosed in patients, which - in the opinion of the echocardiographer - had no clear causative relationship between the lead and TR was observed CIED-related TR defined as a presence of a clear lead interaction, such as lead impingement or adhesion to the valve apparatus.

Based on clinical and echocardiographic findings, patients were evaluated by the local heart teams and, according to established criteria, were either qualified for or disqualified from T-TEER.[8] The primary reason for disqualification was recorded and categorized into following groups: (1) asymptomatic TR; (2) clinical futility (e.g., end-stage heart failure, poor mobility, advanced frailty, or other conditions limiting expected survival); (3) unsuitable TV anatomy (e.g., large coaptation gap, short or tethered leaflets, severe leaflet degeneration, significant lead interference); or (4) insufficient TEE visualization.

If, during the diagnostic process, additional cardiac conditions were identified in referred patients with severe TR, the Heart Team could decide to direct the patient toward another type of intervention instead of T-TEER. Following completion of that intervention, the indication for T-TEER was reassessed, and qualification for the procedure was performed according to the previously established criteria.

In addition to the mentioned reasons for disqualification, for patients in whom the initial interventional treatment led to a reduction of TR severity below the severe threshold, a fifth category — TR grade reduction — was designated as the reason for disqualification.

The protocol of the study was approved by the Ethics Committee of the Medical University of Warsaw (AKBE 179/2023) and endorsed by all participating centers. Data were retrieved from local institutional databases, and all participants provided written informed consent for data collection and analysis.

### Statistical analysis

The statistical analysis was performed using the IBM SPSS Statistics software (version 29.0; IBM, New York, USA). The continuous data were presented either as mean and standard deviation (SD) and compared with student t-test for the normally distributed variables or as a median and interquartile range (IQR) and compared with U Mann-Whitney test for the not-normally distributed variables. The assessment of the distribution of variables was performed using the Shapiro –Wilk test. Categorical variables are presented as a number and percent and compared using the Chi-square test or exact Fisher test. Statistical significance was established at two-sided *P* < 0.05.

## RESULTS

A total of 271 patients with severe TR referred for evaluation of T-TEER were included in the analysis. The mean age of the cohort was 77 ± 9 years, and 58.3% were female. Among them, 113 patients (41.7%) had a cardiac implantable electronic device (CIED) with at least one lead crossing the tricuspid valve.

### Baseline characteristics

Baseline characteristics according to CIED status are summarized in Table 1. Although patients with CIEDs were of similar age compared with those without (79 vs. 76 years, p=0.086), they presented with significantly more comorbidities. The prevalence of diabetes mellitus (35.4% vs. 17.1%, p<0.001) and chronic kidney disease (70.8% vs. 54.4%, p=0.006) was significantly higher among patients with CIEDs. They were also more likely to have a history of CABG (13.3% vs. 4.4%, p=0.009) and were more frequently previously hospitalized for heart failure (78.8% vs. 67.1%, p=0.035).

**Table 1.** Baseline characteristics CIED vs no-CIED.

| Parameter | Overall<br>N=271 | CIED<br>N=113 | No CIED<br>N=158 | P-value |
| --- | --- | --- | --- | --- |
| **Clinical characteristics** |  |  |  |  |
| Female | 158 (58.3) | 60 (53.1) | 98 (62.0) | 0.142 |
| Age | 77.0 (9.0) | 79.0 (11.0) | 76.0 (8.0) | 0.086 |
| NYHA 3/4 | 162 (59.8) | 72 (63.7) | 90 (57.0) | 0.264 |
| Edema | 179 (66.1) | 76 (67.3) | 103 (65.2) | 0.723 |
| Ascites | 54 (19.9) | 27 (23.9) | 27 (17.1) | 0.167 |
| Previous HHF | 195 (72.0) | 89 (78.8) | 106 (67.1) | **0.035** |
| 1 or more HHF in 12 months | 153 (56.5) | 70 (61.9) | 83 (52.5) | 0.123 |
| AF any | 246 (90.8) | 103 (91.2) | 143 (90.5) | 0.857 |
| AF permanent | 187 (69.0) | 77 (68.1) | 110 (69.6) | 0.795 |
| CAD | 106 (39.1) | 49 (43.4) | 57 (36.1) | 0.226 |
| PAD | 24 (8.9) | 10 (8.8) | 14 (8.9) | 0.997 |
| HTN | 192 (70.8) | 79 (69.9) | 113 (71.5) | 0.774 |
| DM | 67 (24.7) | 40 (35.4) | 27 (17.1) | **<0.001** |
| DM insulin | 11 (4.1) | 5 (4.8) | 6 (4.2) | 0.999 |
| CKD | 166 (61.3) | 80 (70.8) | 86 (54.4) | **0.006** |
| COPD/Asthma | 36 (13.3) | 14 (12.4) | 22 (13.9) | 0.714 |
| **Past medical history** |  |  |  |  |
| MI | 55 (20.3) | 29 (25.7) | 26 (16.5) | 0.063 |
| Stroke/TIA | 25 (9.2) | 11 (9.7) | 14 (8.9) | 0.806 |
| PCI | 62 (22.9) | 30 (26.5) | 32 (20.3) | 0.224 |
| CABG | 22 (8.1) | 15 (13.3) | 7 (4.4) | **0.009** |
| TAVI | 13 (4.8) | 5 (4.4) | 8 (5.1) | 0.808 |
| AVR | 20 (7.4) | 6 (5.3) | 14 (8.9) | 0.270 |
| MVR | 25 (9.2) | 10 (8.8) | 15 (9.5) | 0.857 |
| Ablation | 27 (10.0) | 13 (11.5) | 14 (8.9) | 0.485 |
| **Pharmacotherapy** |  |  |  |  |
| Furosemide | 162 (59.8) | 73 (64.6) | 89 (56.3) | 0.171 |
| Torsemide | 174 (64.2) | 78 (69.0) | 95 (60.1) | 0.137 |
| HCTZ | 33 (12.2) | 17 (15.2) | 16 (10.2) | 0.219 |
| Spironolactone | 69 (25.5) | 31 (27.4) | 38 (24.1) | 0.528 |
| Eplerenone | 103 (38.0) | 49 (43.4) | 54 (34.2) | 0.125 |
| BB | 233 (86.0) | 98 (86.7) | 135 (85.4) | 0.764 |
| ACEi | 115 (42.4) | 48 (42.5) | 67 (42.4) | 0.990 |
| ARB | 29 (10.7) | 7 (6.2) | 22 (13.9) | **0.042** |
| ARNI | 15 (5.5) | 13 (11.6) | 2 (1.3) | **<0.001** |
| CCB | 24 (8.9) | 9 (8.0) | 15 (9.5) | 0.662 |
| ASA | 26 (9.6) | 8 (7.1) | 18 (11.4) | 0.235 |
| P2Y12i | 15 (5.5) | 0 (0.0) | 15 (9.5) | **<0.001** |
| VKA | 44 (16.2) | 18 (15.9) | 26 (16.5) | 0.908 |
| NOAC | 180 (66.4) | 80 (70.8) | 100 (63.7) | 0.222 |
| Fractionated heparin | 6 (2.2) | 2 (1.8) | 4 (2.5) | 0.674 |
| Statin | 151 (55.7) | 68 (60.2) | 83 (52.5) | 0.212 |
| SGLT2i | 140 (51.7) | 74 (65.5) | 66 (41.8) | **<0.001** |
| **Laboratory tests** |  |  |  |  |
| Hgb | 12.4 (2.7) | 12.2 (1.9) | 12.2 (2.9) | 0.434 |
| Plt | 177.0 (74.0) | 171.0 (69.0) | 184.0 (79.0) | **0.046** |
| CRP | 1.3 (5.3) | 2.5 (4.4) | 2.2 (5.6) | 0.802 |
| Urea | 58.0 (41.0) | 65.5 (44.6) | 55.0 (33.5) | **0.005** |
| Creatinin | 1.29 (0.58) | 1.40 (0.74) | 1.24 (0.50) | **0.017** |
| eGFR | 46.0 (24.3) | 44.9 (16.1) | 48.0 (24.0) | 0.078 |
| AST | 31.0 (12.0) | 31.0 (12.0) | 30.0 (12.0) | 0.140 |
| ALT | 21.0 (13.0) | 20.0 (13.0) | 22.0 (12.0) | 0.673 |
| INR | 1.31 (0.57) | 1.37 (0.80) | 1.25 (0.60) | 0.111 |
| Bilirubin | 0.92 (0.87) | 1.00 (1.04) | 0.84 (0.81) | **0.028** |
| NT-proBNP | 1732.0<br>(2443.0) | 2033.0 (2904.0) | 1573.0 (1907.0) | **0.046** |
| Na | 139.3 (4.2) | 139.3 (4.1) | 139.4 (5.0) | 0.369 |
| **Echocardiographic characterization** |  |  |  |  |
| MR |  |  |  | 0.640 |
| 0 | 21 (7.7) | 9 (8.4) | 12 (7.9) |  |
| 1 | 107 (39.5) | 42 (39.3) | 65 (43.0) |  |
| 2 | 72 (26.6) | 35 (32.7) | 37 (24.5) |  |
| 3 | 43 (15.9) | 15 (14.0) | 28 (18.5) |  |
| 4 | 15 (5.5) | 6 (5.6) | 9 (6.0) |  |
| Missing data | 13 (4.8) | 6 (5.3) | 7 (4.4) |  |
| TR baseline |  |  |  | 0.099 |
| 3 | 80 (29.5) | 31 (27.4) | 49 (31.0) |  |
| 4 | 102 (37.6) | 36 (31.9) | 66 (41.8) |  |
| 5 | 89 (32.8) | 46 (40.7) | 43 (27.2) |  |
| LVDd | 4.9 (1.2) | 5.1 (1.2) | 4.9 (0.8) | **0.003** |
| IVSd | 1.0 (0.3) | 1.0 (0.3) | 1.0 (0.3) | 0.250 |
| PWTd | 1.0 (0.2) | 1.0 (0.2) | 1.0 (0.2) | 0.244 |
| RVOT | 3.7 (1.0) | 3.8 (1.1) | 3.7 (1.0) | 0.259 |
| LA | 5.2 (1.0) | 5.2 (0.9) | 5.1 (1.0) | 0.364 |
| EF | 55.0 (15.0) | 50.5 (20.0) | 57.0 (12.0) | **<0.001** |
| RAA | 34.8 (14.0) | 35.0 (14.0) | 34.0 (14.4) | 0.269 |
| RVIT | 5.0 (0.8) | 5.1 (0.7) | 4.9 (0.8) | 0.088 |
| TAPSE | 17.0 (6.0) | 16.3 (4.4) | 18.0 (4.9) | **0.007** |
| MR ERO | 0.25 (0.16) | 0.23 (0.13) | 0.26 (0.24) | 0.514 |
| MR Vol | 37.0 (33.0) | 33.5 (27.0) | 41.7 (21.3) | 0.594 |
| TR ERO | 0.62 (0.34) | 0.70 (0.38) | 0.60 (0.30) | **0.012** |
| TR Vol | 60.0 (30.0) | 62.0 (36.0) | 59.1 (19.0) | 0.246 |

ARB – angiotensin II receptor blocker; ARNI – angiotensin receptor–neprilysin inhibitor;
AVR – aortic valve replacement; BB – beta-blocker; CABG – coronary artery bypass grafting;
CAD – coronary artery disease; CCB – calcium channel blocker; CKD – chronic kidney
disease; COPD – chronic obstructive pulmonary disease; CRP – C-reactive protein; DM –
diabetes mellitus; eGFR – estimated glomerular filtration rate; HCTZ – hydrochlorothiazide;
HHF – hospitalization for heart failure; HTN – hypertension; IVSd – interventricular septal
diameter (diastolic); LA – left atrium; LVDd – left ventricular diastolic diameter; MI –
myocardial infarction; MR – mitral regurgitation; MR ERO – mitral regurgitant effective
regurgitant orifice; MR Vol – mitral regurgitant volume; MVR – mitral valve replacement; Na

Laboratory and imaging data further revealed decreased renal function and a more advanced associated organ dysfunction in the CIED group: higher urea concentration (65.5 vs. 55.0 mg/dL, p=0.005), higher creatinine concentration (1.40 vs. 1.24 mg/dL, p=0.017), higher bilirubin concentration (1.00 vs. 0.84 mg/dL, p=0.028), lower platelet count (171 vs. 184 × 10^3/µL, p=0.046), and higher NT-proBNP levels (2033 vs. 1573 pg/mL, p=0.046). Echocardiography demonstrated lower left ventricular ejection fraction (50.5% vs. 57.0%, p<0.001), lower TAPSE (16.3 vs. 18.0 mm, p=0.007), larger LV end-diastolic diameter (5.1 vs. 4.9 cm, p=0.003), and greater TR effective regurgitant orifice (0.70 vs. 0.60 cm², p=0.012) in patients with CIEDs.

Medication use also differed between groups. Patients with CIEDs were more frequently treated with ARNI (11.6% vs. 1.3%, p<0.001) and SGLT2 inhibitors (65.5% vs. 41.8%, p<0.001).

### Qualification for T-TEER

A detailed patient flow and qualification process are summarized in Figure 1. Overall, 153 patients (56.5%) were qualified for T-TEER. The qualification rate did not differ significantly between patients with and without CIEDs (54.0% vs. 58.3%, p=0.487). Among the 113 disqualified patients, the most frequent reasons were unsuitable anatomy (31.0%), clinical futility (29.2%), and asymptomatic disease (23.0%), while insufficient TEE visualization (5.3%) and TR grade reduction (11.5%) were less common (Table 2, Figure 2). The distribution of disqualification reasons was similar in patients with and without CIEDs.

**Figure 1.**
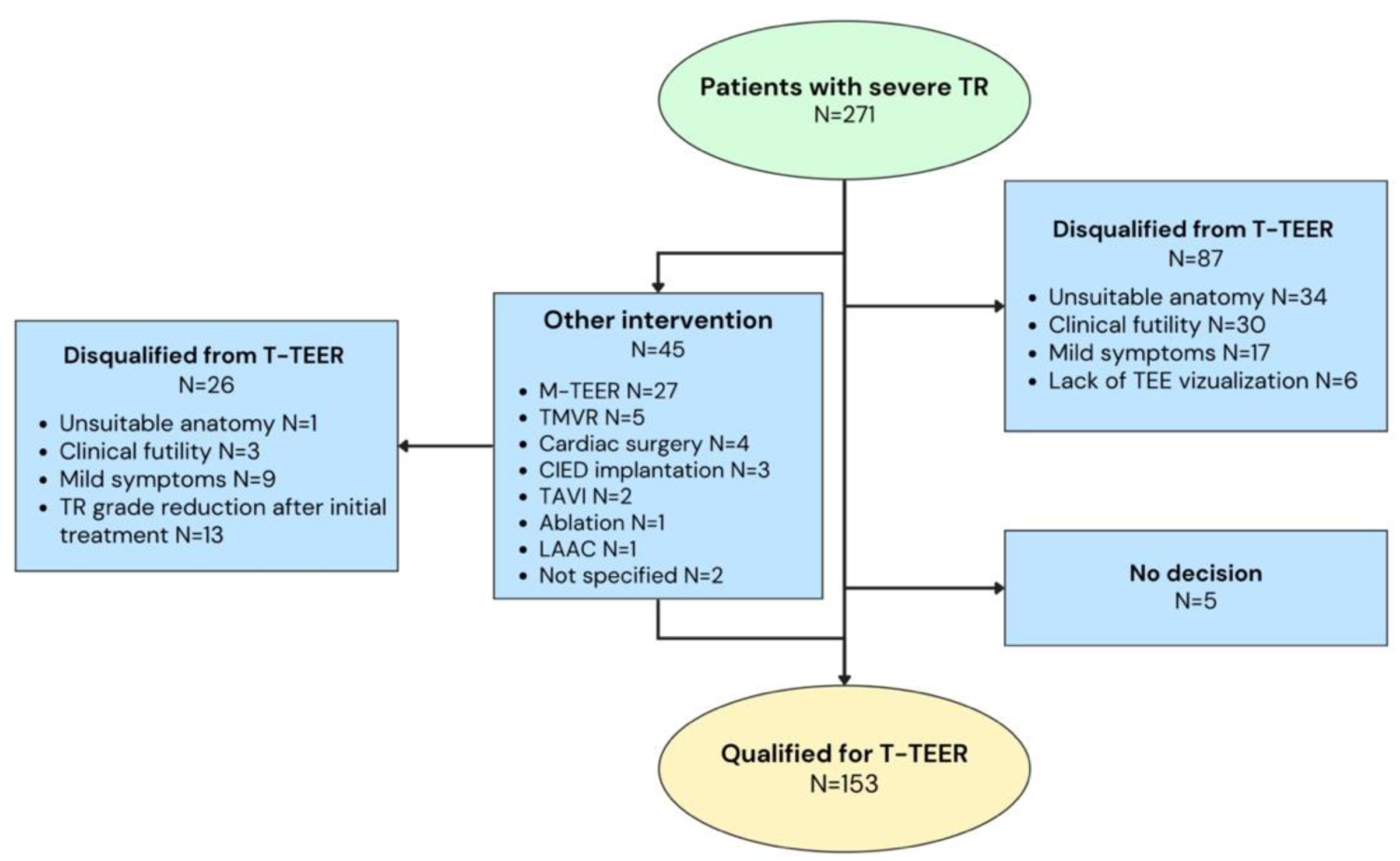
Patient flow and qualification outcomes for T-TEER.

**Figure 2.**
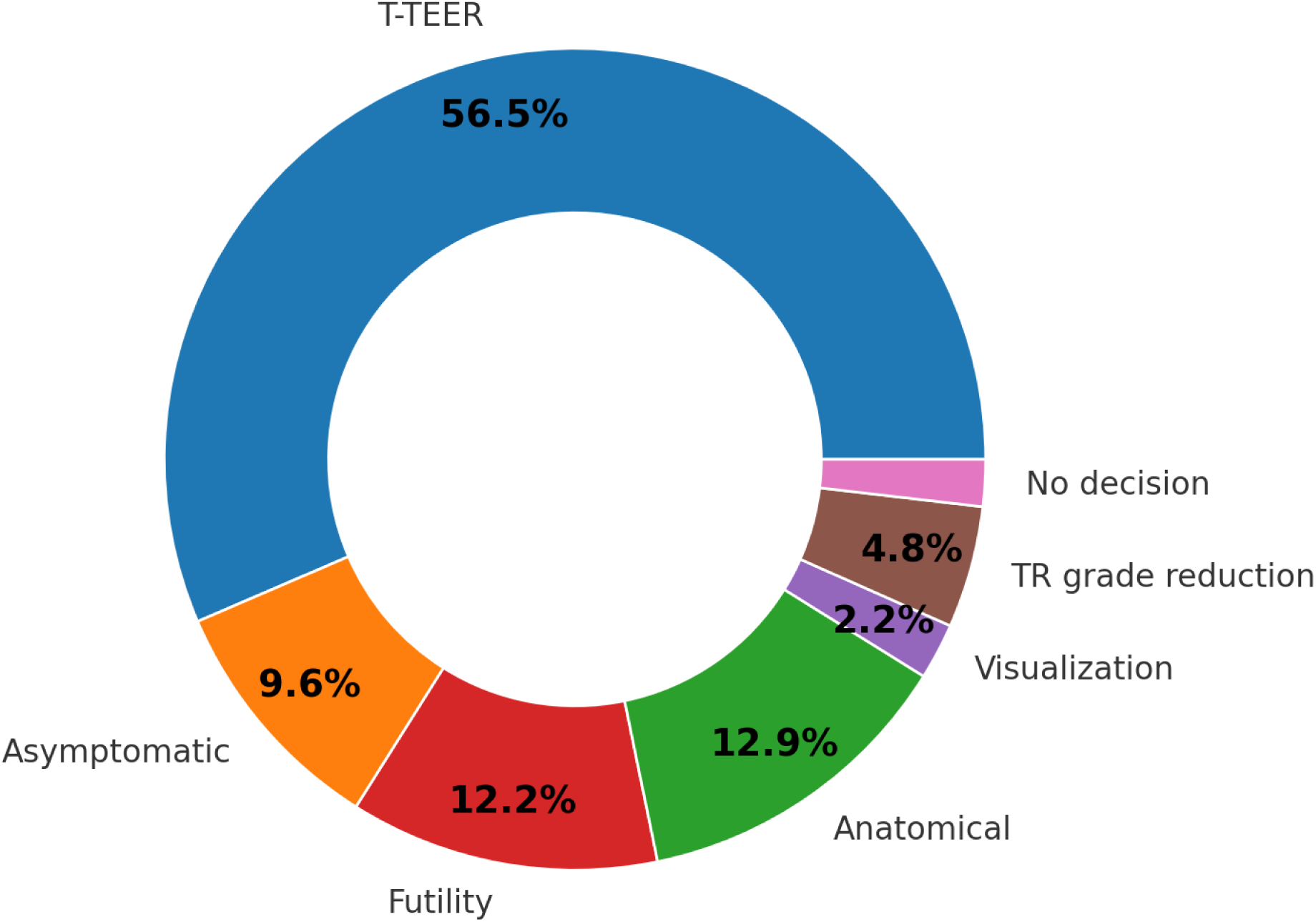
Final qualification outcomes among patients referred for T-TEER.

**Table 2.**
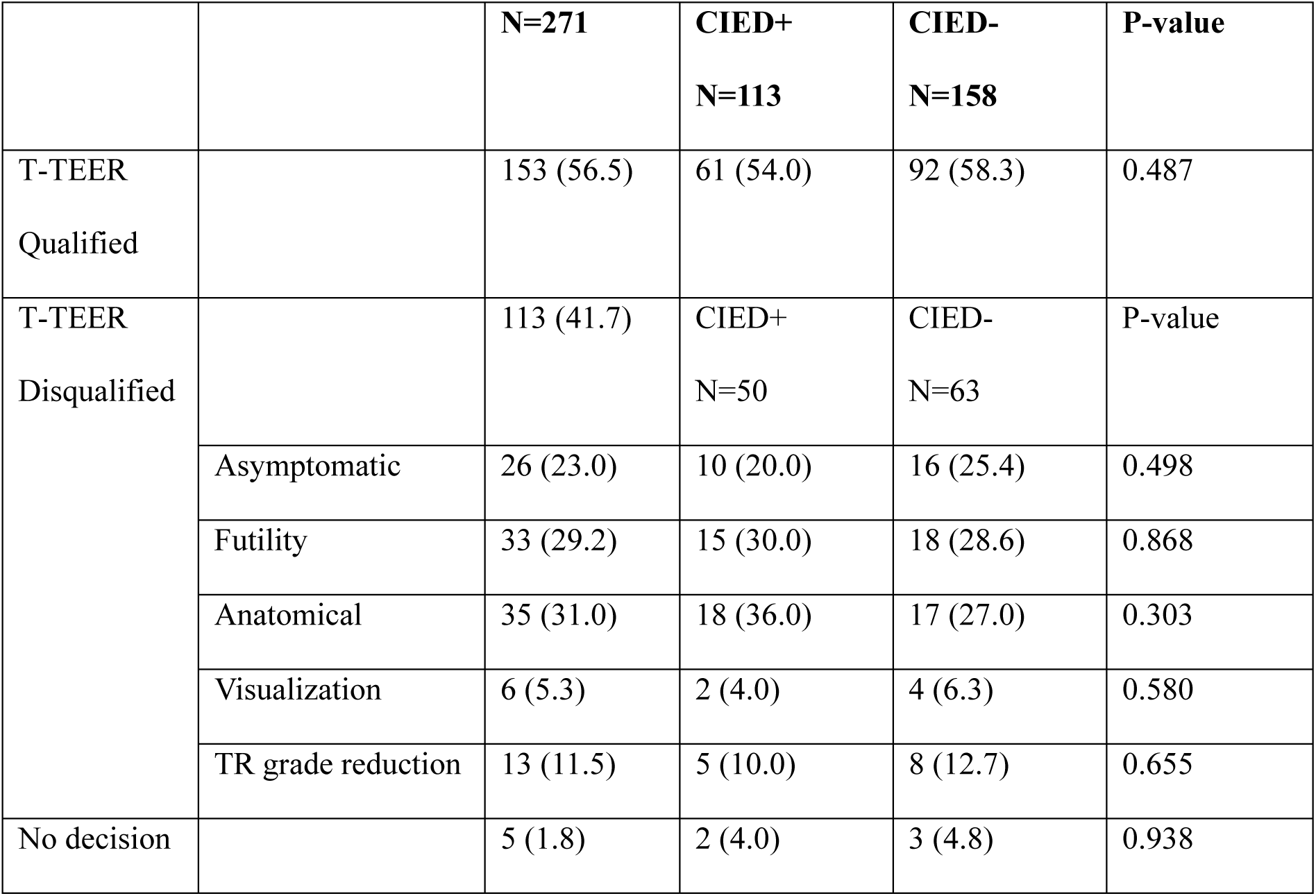
Decision regarding treatment and reasons of T-TEER disqualification in patients with and without CIED.

### CIED-related vs. CIED-associated TR

Among the 113 patients with CIEDs, 37 (32.7%) were classified as having CIED-related TR and 76 (67.3%) as CIED-associated TR. Baseline characteristics are presented in Table 3. Compared with CIED-associated TR, patients with CIED-related TR more frequently presented with peripheral edema (81.1% vs. 60.5%, p=0.029) and ascites (40.5% vs. 15.8%, p=0.004). Permanent atrial fibrillation was less common in CIED-related patients (54.1% vs. 75.0%, p=0.025), while the use of eplerenone (56.8% vs. 36.8%, p=0.045) and SGLT2 inhibitors (81.1% vs. 57.9%, p=0.015) was more frequent.

**Table 3.** CIED-related vs CIED-associated.

| **Parameter** | **CIED-related**<br>**N=37** | **CIED-associated**<br>**N=76** | **P-value** |
| --- | --- | --- | --- |
| **Clinical characteristics** |  |  |  |
| Female | 19 (51.4) | 41 (53.9) | 0.795 |
| Age | 77.6 (8.7) | 79.0 (11.0) | 0.849 |
| NYHA 3/4 | 27 (73.0) | 45 (59.2) | 0.153 |
| Edema | 30 (81.1) | 46 (60.5) | **0.029** |
| Ascites | 15 (40.5) | 12 (15.8) | **0.004** |
| Previous HHF | 29 (78.4) | 60 (78.9) | 0.945 |
| 1 or more HHFin 12 months | 23 (62.2) | 47 (61.8) | 0.974 |
| AF any | 34 (91.9) | 69 (90.8) | 0.846 |
| AF permanent | 20 (54.1) | 57 (75.0) | **0.025** |
| CAD | 15 (40.5) | 34 (44.7) | 0.673 |
| PAD | 3 (8.1) | 7 (9.2) | 0.846 |
| HTN | 24 (64.9) | 55 (72.4) | 0.414 |
| DM | 10 (27.0) | 30 (39.5) | 0.194 |
| DM insulin | 0 (0.0) | 5 (7.2) | 0.162 |
| CKD | 28 (75.7) | 52 (68.4) | 0.426 |
| COPD/Asthma | 5 (11.3) | 9 (11.8) | 0.770 |
| **Past medical history** |  |  |  |
| MI | 8 (21.6) | 21 (27.6) | 0.492 |
| Stroke/TIA | 3 (8.1) | 8 (10.5) | 0.684 |
| PCI | 6 (16.2) | 24 (31.6) | 0.083 |
| CABG | 5 (13.5) | 10 (13.2) | 0.958 |
| TAVI | 1 (2.7) | 4 (5.3) | 0.535 |
| AVR | 0 (0.0) | 6 (7.9) | 0.175 |
| MVR | 2 (5.4) | 8 (10.5) | 0.368 |
| Ablation | 5 (13.5) | 8 (10.5) | 0.640 |
| **Pharmacotherapy** |  |  |  |
| Furosemide | 26 (70.3) | 47 (61.8) | 0.379 |
| Torsemide | 27 (73.0) | 51 (67.1) | 0.676 |
| HCTZ | 6 (16.7) | 11 (14.5) | 0.763 |
| Spironolactone | 7 (18.9) | 24 (31.6) | 0.157 |
| Eplerenone | 21 (56.8) | 28 (36.8) | **0.045** |
| BB | 33 (89.2) | 65 (85.5) | 0.590 |
| ACEi | 17 (45.9) | 31 (40.8) | 0.603 |
| ARB | 2 (5.4) | 5 (6.6) | 0.808 |
| ARNI | 1 (2.8) | 12 (15.8) | 0.058 |
| CCB | 4 (10.8) | 5 (6.6) | 0.436 |
| ASA | 2 (5.4) | 6 (7.9) | 0.628 |
| P2Y12i | 0 | 0 | - |
| VKA | 7 (18.9) | 11 (14.5) | 0.545 |
| NOAC | 26 (70.3) | 54 (71.1) | 0.932 |
| Fractionated heparin | 2 (5.4) | 0 (0.0) | 0.105 |
| Statin | 27 (73.0) | 41 (53.9) | 0.053 |
| SGLT2i | 30 (81.1) | 44 (57.9) | **0.015** |
| **Laboratory tests** |  |  |  |
| Hgb | 12.4 (1.7) | 12.1 (2.0) | 0.465 |
| Plt | 180.0 (85.0) | 166.5 (72.0) | 0.159 |
| CRP | 2.2 (6.7) | 2.5 (4.2) | 0.719 |
| Urea | 72.4 (33.1) | 63.0 (43.4) | 0.865 |
| Creatinin | 1.42 (0.53) | 1.33 (0.89) | 0.525 |
| eGFR | 43.6 (14.1) | 45.6 (17.0) | 0.537 |
| AST | 31.0 (12.0) | 31.5 (13.0) | 0.849 |
| ALT | 20.0 (10.0) | 19.5 (15.0) | 0.839 |
| INR | 1.36 (0.57) | 1.37 (0.90) | 0.750 |
| Bilirubin | 0.99 (0.78) | 1.0 (1.31) | 0.313 |
| NT-proBNP | 2467.5 (4158.3) | 1875.0 (2694.0) | 0.309 |
| Na | 139.7 (4.3) | 139.0 (4.1) | 0.753 |
| **Echocardiographic characterization** |  |  |  |
| MR |  |  | 0.377 |
| 0 | 2 (5.6) | 7 (9.9) |  |
| 1 | 16 (44.4) | 26 (36.6) |  |
| 2 | 12(33.3) | 23 (32.4) |  |
| 3 | 6 (16.7) | 9 (12.7) |  |
| 4 | 0 (0.0) | 6 (8.5) |  |
| Missing data | 1 (2.7) | 5 (6.6) |  |
| TR baseline |  |  | 0.006 |
| 3 | 4 (10.8) | 27 (35.5) |  |
| 4 | 11 (29.7) | 25 (32.9) |  |
| 5 | 22 (59.5) | 24 (31.6) |  |
| LVDd | 5.2 (0.9) | 5.2 (1.2) | 0.891 |
| IVSd | 1.0 (0.4) | 1.1 (0.3) | 0.476 |
| PWTd | 1.0 (0.3) | 1.0 (0.2) | 0.298 |
| RVOT | 4.1 (0.8) | 3.7 (1.0) | 0.066 |
| LA | 5.2 (1.1) | 5.3 (0.8) | 0.654 |
| EF | 50.4 (11.8) | 50.0 (18.0) | 0.583 |
| RAA | 35.0 (21.3) | 35.0 (13.0) | 0.800 |
| RVIT | 4.9 (0.8) | 5.2 (0.7) | 0.243 |
| TAPSE | 16.7 (4.8) | 16.1 (4.2) | 0.490 |
| MR ERO | 0.23 (0.11) | 0.21 (0.15) | 0.897 |
| MR Vol | 29.5 (31.0) | 37.0 (26.0) | 0.716 |
| TR ERO | 0.90 (0.34) | 0.60 (0.35) | **0.017** |
| TR Vol | 66.6 (26.4) | 58.0 (37.0) | 0.428 |

Echocardiographic evaluation showed that patients with CIED-related TR had a higher prevalence of torrential TR (59.5% vs. 31.6%, p=0.006) and a larger TR effective regurgitant orifice (0.90 vs. 0.60 cm², p=0.017) as shown in Figure 3

**Figure 3.**
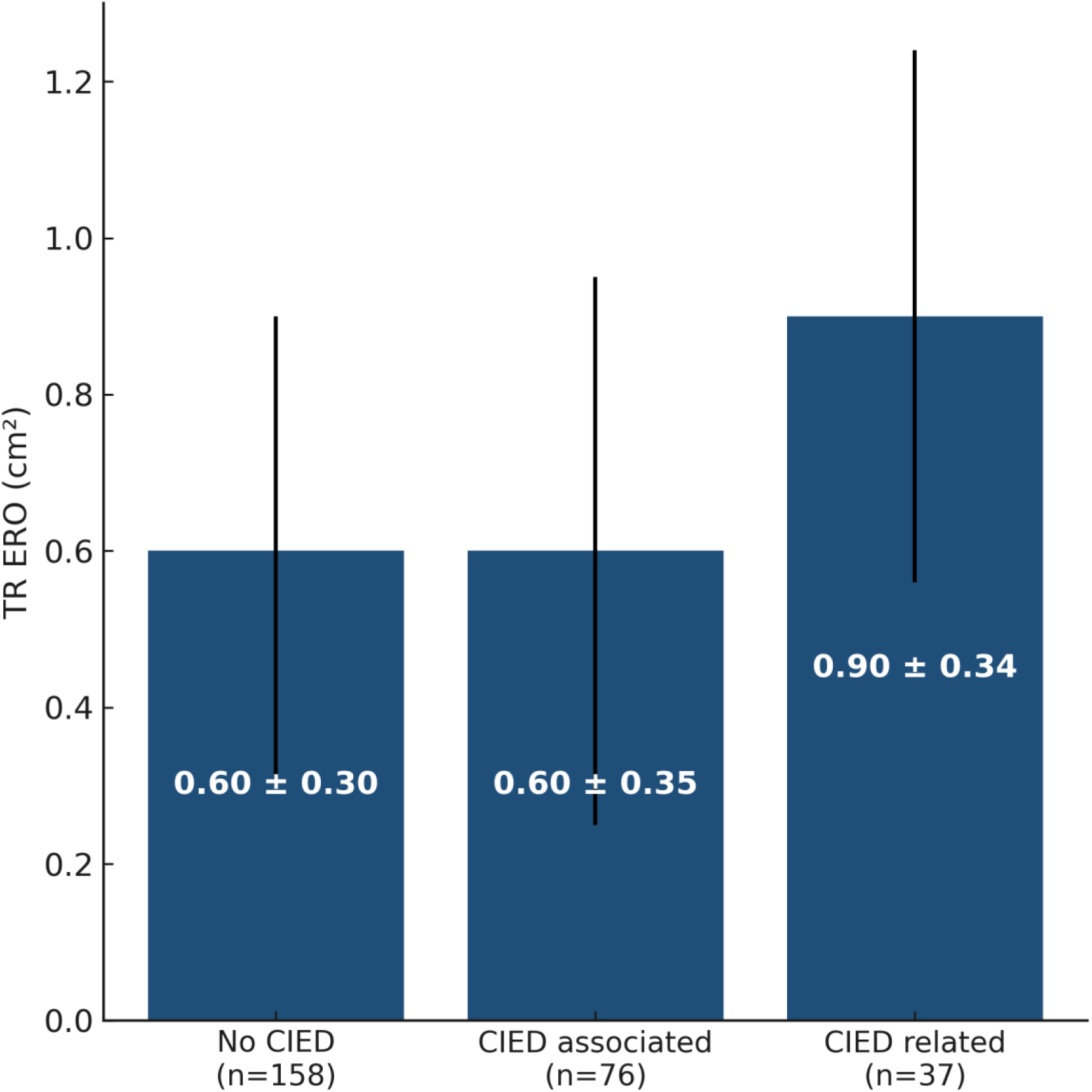
Tricuspid regurgitation effective orifice area according to CIED status. CIED related vs. CIED associated and CIED related vs. no CIED - p<0,05; CIED associated vs. no CIED – p=NS

### T-TEER qualification according to type of CIED interaction

The rate of T-TEER qualification did not differ significantly between patients with CIED-related and CIED-associated TR (59.5% vs. 52.7%, p=0.500; Table 4). Similarly, the distribution of reasons for disqualification was comparable between the two subgroups, with anatomical factors representing the leading cause in both groups.

**Table 4.** Decision regarding treatment and reasons of T-TEER disqualification in patients with CIED related and CIED associated TR.

|  |  | **CIED +**<br>**N=113** | **CIED related**<br>**N=37** | **CIED**<br>**associated**<br>**N=74** | **P-value** |
| --- | --- | --- | --- | --- | --- |
| T-TEER<br>Qualified |  | 61 (54.0) | 22 (59.5) | 39 (52.7) | 0.500 |
| T-TEER<br>Disqualified |  | CIED+<br>N=50 | CIED related<br>N=15 | CIED<br>associated<br>N=35 | P-value |
|  | Asymptomatic | 10 (20.0) | 2 (13.3) | 8 (22.9) | 0.440 |
|  | Futility | 15 (30.0) | 4 (26.7) | 11 (31.4) | 0.736 |
|  | Anatomical | 18 (36.0) | 6 (40.0) | 12 (34.3) | 0.699 |
|  | Visualization | 2 (4.0) | 1 (6.7) | 1 (2.9) | 0.999 |
|  | TR grade<br>reduction | 5 (10.0) | 2 (13.3) | 3 (8.6) | 0.607 |
| No decision |  | 2 (1.8) | 0 (0.0) | 2 (2.7) | 0.999 |

## DISCUSSION

The present study provides the first comprehensive evaluation of the impact of CIEDs on the T-TEER qualification process among patients with severe TR referred to tertiary centers. The major findings expand current knowledge in this field and could be summarized as follows: First, the prevalence of CIEDs in our cohort was higher than previously reported. Prior studies on tricuspid T-TEER have shown considerable variation in the reported frequency of CIEDs. For instance, in the original population of randomized TRILUMINATE Pivotal Trial, only 16% of patients in the T-TEER group had a CIED.[9] In the recently published bRIGHT EU PAS registry, which specifically addressed the history of CIEDs implantation, patients with a TV-crossing lead represented 21.6% of the study population.[4] Furthermore, a recent scientific statement reported that the prevalence of CIEDs among patients undergoing transcatheter tricuspid valve interventions ranges from 11.8% to 36%.[1] In the present study, 41.7% of patients referred for TR treatment carried a CIED, which, to our knowledge, represents the highest prevalence reported to date and highlights the clinical relevance of this problem in a real-world population.

Second, among patients with CIEDs, nearly one in three were classified as having CIED-related tricuspid regurgitation, caused by direct mechanical interaction between the lead and the tricuspid valve apparatus. CIED-related TR has recently been recognized as a distinct clinical entity, clearly differentiated from CIED-associated TR, in which the TV-crossing lead acts as a bystander rather than the primary cause of regurgitation.[2] To date, there are no uniform diagnostic criteria allowing for an unequivocal definition of this phenomenon, and its identification relies largely on subjective echocardiographic assessment. Consequently, the true prevalence of lead-related TR remains uncertain. In a recent prospective cohort study, CIED-related TR represented approximately 5% of all severe TR cases, [10] whereas in our study this condition was observed in 13.6% of patients with severe TR, highlighting its potentially greater impact in a real-world referral population.

Third, patients with CIEDs carried a greater overall comorbidity burden compared with those without CIEDs. The CIED group was characterized by significantly higher rates of diabetes mellitus, chronic kidney disease, and prior coronary revascularization, as well as worse laboratory and echocardiographic parameters. Patients with CIEDs, and particularly those with CIED-related TR, received more intensive pharmacological treatment, which may reflect a more advanced stage of heart failure in this subgroup. These findings suggest that the presence of a CIED frequently identifies a more advanced and complex patient profile. Within the CIED cohort, patients with CIED-related TR were more symptomatic, showing higher rates of peripheral edema and ascites, and more frequently exhibited torrential regurgitation with larger effective regurgitant orifice area compared with CIED-associated TR patients. This subgroup therefore represents a particularly severe clinical phenotype.

Fourth, in the present study, approximately half of the referred patients were qualified for T-TEER. Until now, only a few published studies focused on the selection of patients for invasive TR treatment. In a retrospective analysis involving 547 patients from three centers, 196 (35.8%) patients were qualified for T-TEER, while a total of 136 (24.9%) patients were referred to other transcatheter therapeutic modalities mainly direct annuloplasty and in minority of cases transcatheter valve implantation.[11] In another retrospective study involving patients evaluated for tricuspid interventions, anatomical feasibility for T-TEER and transcatheter tricuspid valve implantation were analyzed. Among 491 patients assessed for T-TEER, 157 (32.0%) were found to have unfavorable anatomy for percutaneous valve repair attempt.[12] In our cohort, the most common reason for disqualification from T-TEER was unfavorable valve anatomy, which is consistent with findings from previous studies and highlights the clinical need for broader implementation of alternative transcatheter tricuspid interventions.

Finally, no differences were observed in T-TEER qualification rates or reasons for disqualification between patients with and without CIEDs. Moreover, the CIED-related and CIED-associated subgroups showed comparable qualification rates. To date, no studies have systematically addressed this issue. In a study by Tanaka et al., the highest prevalence of CIEDs was reported among patients classified as unfavorable for T-TEER (44.6%), compared with a significantly lower prevalence in the favorable (5%) and feasible (28.5%) groups.[12] However, that study did not analyze final treatment decisions or procedural qualification, but rather described the eligibility of patients for various transcatheter treatment modalities. Conversely, the absence of differences in qualification rates observed in our study may reflect the limited impact of CIED presence on procedural feasibility and T-TEER outcomes, as also suggested by previous reports.

## LIMITATIONS

This study has several limitations that should be acknowledged. First, its retrospective design carries the risk of selection bias and limits the ability to establish causal relationships. Second, echocardiographic assessment of TR severity, mechanism, and lead–valve interaction was not performed by an independent core laboratory but relied on evaluations conducted at participating centers. This approach may have introduced some variability in image interpretation. Third, decisions regarding T-TEER qualification were made by local heart teams, which could have led to differences in patient selection criteria between centers. On the other hand, these features also reflect a real-world clinical practice, offering a representative overview of referral patterns and qualification processes for transcatheter tricuspid interventions.

## CONCLUSIONS

CIED carriers referred to tertiary centers with severe TR represent a more complex and symptomatic population compared with non-CIED patients. Nevertheless, the ultimate decision to proceed with T-TEER is not determined by the CIED status.

Furthermore, the high prevalence of CIEDs and the considerable proportion of patients with lead-related TR observed in this study underscore the growing clinical relevance of this issue in contemporary practice. Our findings highlight the need for prospective, multicenter studies to validate these observations and to further refine the treatment qualification process in patients with severe TR and concomitant CIEDs.

## Data Availability

The data that support the findings of this study are available from the corresponding author upon reasonable request. Due to privacy and ethical restrictions, individual patient data are not publicly available.

## Notes

**Conflict of interest**: All authors declare no conflict of interest regarding the contents of this article.

### Competing Interest Statement

Adam Rdzanek: consulting fees and speaker honoraria from Edwards Lifesciences Piotr Scis?o: consulting fees and speaker honoraria from Edwards Lifesciences Francesco Maisano: Grant and/or Research Institutional Support from Abbott, Medtronic, Edwards Lifesciences, Biotronik, Boston Scientific Corporation, NVT, Terumo, Venus; Roche Consulting fees, Honoraria personal and Institutional from Abbott, Medtronic, Edwards Lifesciences, Xeltis, Cardiovalve, Occlufit, Simulands, Mtex, Venus, Squadra, Meril, Croivalve; Royalty Income/IP Rights from Edwards Lifesciences and is Shareholder (including share options) of Magenta, Transseptalsolutions, 4Tech.

### Clinical Trial

NCT06838611

### Funding Statement

No external funding was received for this work.

### Author Declarations

The study was conducted in accordance with the Declaration of Helsinki and approved by the Ethics Committee of the Medical University of Warsaw (AKBE 179/2023)

